# Daily Stress and Heart Rate Variability Among Mindfulness Meditation Practitioners: An mHealth Observational Study

**DOI:** 10.1101/2025.08.16.25333623

**Authors:** Jo Takezawa, Shixian Geng, Masahiro Fujino, Mika Miyake, Kazutoshi Sasahara, Koji Yatani, Atsushi Niida

**Affiliations:** Interactive Intelligent Systems Laboratory, The University of Tokyo; Human Information Science Laboratory, NTT Communication Science Laboratories; Garmin Health, Garmin Japan Ltd.; Department of Innovation Science, School of Environment and Society, Institute of Science Tokyo; Laboratory of Molecular Medicine, Human Genome Center, The Institute of Medical Science, The University of Tokyo

**Keywords:** Mindfulness meditation, Stress reduction, Heart rate variability, Experience sampling method, Smartwatch

## Abstract

**Background:** Mindfulness meditation has been reported to reduce stress and enhance well-being. However, its effects on heart rate variability (HRV) – a physiological marker of stress – remain underexplored.

**Objective:** To examine how meditation practice is associated with subjective stress, HRV, and their interaction, using mobile health (mHealth) technologies.

**Methods:** This three-week observational study included 90 participants: 19 meditation practitioners, 32 recreational runners, and 39 individuals without regular meditation or exercise habits. HRV was continuously recorded using Garmin smartwatches. Subjective stress and activity were assessed three times daily via a smartphone-based experience sampling method (ESM), yielding 4,557 responses; 632 meditation sessions were time-logged; questionnaires were administered at the end of the study.

**Results:** Stress (PSS) was lower in meditation and running groups vs. controls (Kruskal–Wallis *P* = .024; adjusted Wilcoxon *P* = .048 and .040). RMSSD was higher in runners vs. controls (*P* < .001) but not different between meditation and controls. Higher perceived stress was associated with concurrent RMSSD reduction of −2.24 ms (95% CI −3.97 to −0.26), steeper in runners (−3.94 ms; 95% CI −7.04 to −0.74). During meditation, RMSSD increased by +4.68 ms (95% CI 2.96 to 6.38) and remained elevated for at least 30 minutes post-practice.

**Conclusions:** Although daily-life HRV among meditation practitioners was not elevated overall, their ability to increase HRV at will with a prolonged residual effect may relate to stress reduction and warrants further investigation.

**Trial Registration:** Not applicable.

## Introduction

Mindfulness refers to a psychological state characterized by present-moment awareness and non-judgmental acceptance of one’s experiences [1]. Although originally rooted in Buddhist traditions, mindfulness meditation (hereafter referred to simply as meditation) has been redefined as a secular mental practice aimed at cultivating mindfulness, and is now widely adopted in non-religious contexts for its potential to reduce stress and enhance well-being [2–3]. Numerous studies have demonstrated the effectiveness of meditation in reducing perceived stress through self-report measures [4–6]. However, these studies have primarily assessed stress levels in constrained laboratory settings or before and after fixed intervention periods, leaving the impact of daily meditation practice on real-world stress largely unexplored.

In addition to retrospective questionnaires, subjective stress levels can be assessed using the experience sampling method (ESM), also known as ecological momentary assessment (EMA) [7–8]. While questionnaires are suitable for capturing general trends in perceived stress, ESM enables participants to report their thoughts, emotions, and behaviors multiple times per day in real time, within their natural environments. This approach enables the simultaneous collection of contextual information, facilitating the examination of temporal relationships between stress and other variables in everyday life. Recently, ESM has been employed in meditation researches to evaluate the stress reduction effects in real-world contexts [9–10].

Heart rate variability (HRV) is widely used in stress research as a physiological marker of stress. HRV refers to fluctuations in the time intervals between successive heartbeats and serves as an index of autonomic nervous system balance. Elevated stress levels are typically associated with reduced HRV [11–12]. While HRV is traditionally assessed using electrocardiography (ECG), recent advances in mobile health (mHealth) technologies have facilitated the widespread use of consumer-grade wearable devices—such as smartwatches and fitness trackers—that utilize photoplethysmography (PPG) for physiological monitoring [13–14]. Although PPG-based devices are more susceptible to noise than ECG, they offer advantages such as portability, extended battery life, and the ability to perform long-term HRV monitoring in real-world environments. Moreover, their affordability and widespread adoption facilitate large-scale population-based studies [15].

Laboratory-based studies have consistently demonstrated an inverse association between HRV and perceived stress, particularly under experimentally induced stress conditions. However, findings in everyday life settings have been inconsistent [16–19]. More recently, studies combining ESM with PPG-based wearable devices for HRV monitoring have begun to clarify this relationship in real-world contexts; although the observed effects are generally modest, elevated perceived stress is significantly associated with reduced HRV in real time [20].

Although transient increases in HRV during meditation have been consistently reported in laboratory settings, whether long-term meditation practice yields sustained enhancements in HRV remains uncertain, as findings from intervention studies have been mixed [21–25]. Moreover, to date, there has been little to no observational research examining HRV patterns during daily life or during meditation practice among experienced meditation practitioner. In contrast, regular physical activity—like meditation—is well known to confer stress-reducing benefits [26–27], and athletes have been consistently shown to exhibit elevated HRV levels in everyday life compared to non-athletic individuals [28–29].

In summary, despite growing evidence linking meditation to stress reduction and HRV elevation, the lack of real-world monitoring has limited our understanding of how these relationships unfold in daily life. We designed an observational study to investigate the relationship between stress and HRV in experienced meditation practitioners, integrating smartwatch-based HRV monitoring and ESM. The study is conducted on 90 subjects consisting of three groups: 19 meditation practitioners (meditation group), 32 recreational runners as an active control group characterized by lower stress and higher HRV (running group), and 39 individuals with neither a regular meditation nor exercise routine as a passive control group (control group). Over a period of three weeks, participants continuously wore Garmin smartwatches to monitor HRV. In parallel, smartphone-based ESM was used to assess momentary perceived stress and activity states throughout daily life. At the end of the monitoring period, participants completed a web-based questionnaire evaluating subjective stress and related factors. Furthermore, for the meditation group, the timing of each daily meditation practice was logged via the smartphone app, allowing the capture of HRV dynamics during and after practice. These multimodal datasets were integrated to address the following key questions.

1. Do meditation practitioners exhibit lower levels of perceived stress in daily life, consistent with previous findings?
2. Are HRV levels in daily life elevated among meditation practitioners compared to control participants?
3. Do real-time associations of stress with activity states and HRV differ between meditation practitioners and control participants?
4. During regular meditation practices, do meditation practitioners show increases in HRV, as reported by laboratory-based studies, and if so, how long are such increases sustained after meditation practices?

Taken together, this study seeks to quantitatively assess how meditation practice is associated with subjective stress, HRV, and their interaction in real-world settings, leveraging mobile health technologies to extend beyond the constraints of controlled laboratory environments (Figure 1).

**Figure 1:**
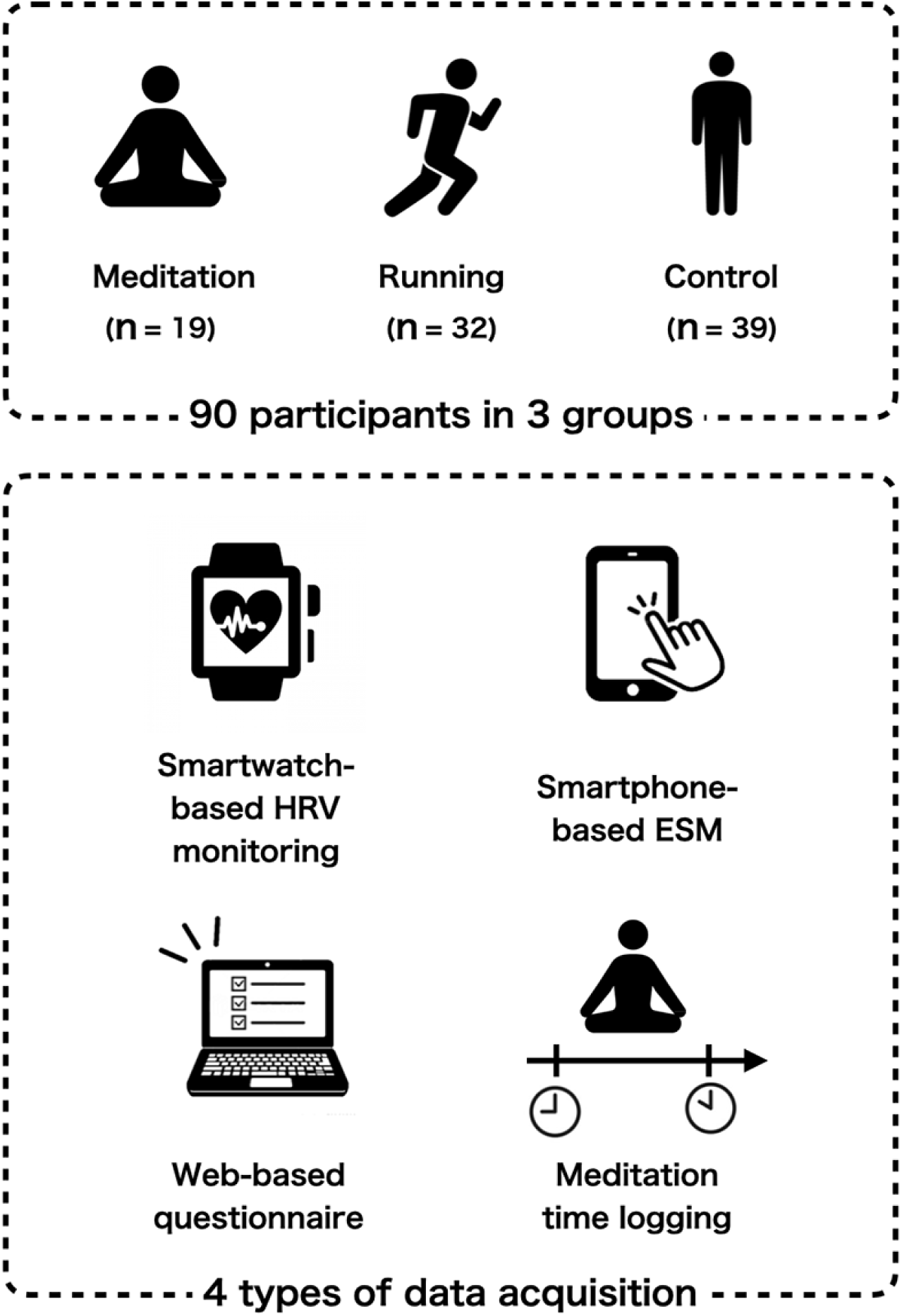
Graphical Summary of our study design. Data were obtained from 90 participants divided into three groups: 19 meditation practitioners (meditation group), 32 recreational runners (runner group), and 39 individuals with neither regular meditation nor exercise routines (control group). Over a three-week period, participants continuously wore Garmin smartwatches to monitor HRV. In parallel, a smartphone-based ESM was used to assess momentary stress and activity in daily life. At the end of the study, participants completed a web-based questionnaire on subjective stress and related psychological variables. For the meditation group, daily practice times were logged via a smartphone app to enable fine-grained analysis of HRV dynamics during and after meditation.

## Methods

### Participants and Recruitment

We recruited healthy adults aged 18 years and older who did not receive medical treatment or take regular medication. We grouped participants based on the following criteria:

1. Meditation Group Participants must have at least one year of experience in secular mindfulness meditation (e.g., MBSR, Mindfulness-Based Cognitive Therapy [MBCT]) or meditation based on traditional schools of Buddhism (e.g., Theravada, Zen). They must practice meditation for at least 20 minutes a day, five days a week, and meet at least one of the following criteria:
  (a) Participation in an intensive meditation retreat of at least one week.
  (b) Completion of a teacher training course in the Method of the Clinical Meditation [30].
  (c) Completion of an eight-week mindfulness program (e.g., MBSR, MBCT).
2. Running Group Participants must have no regular practice of meditation or yoga, no participation in meditation courses (retreats or multi-week programs), and must run for at least 30 minutes a day, four days a week, for over a year.
3. Control Group Participants must have no regular practice of meditation or yoga, no participation in meditation courses, and exercise no more than two days a week for a maximum of 30 minutes per day.

All participants were required to own an iPhone, as data collection and experience sampling were conducted using UTracker, an in-house iOS app. The app is compatible with Garmin watch models Vivosmart 4, ForeAthlete 245/245M, and ForeAthlete 745. We provided Vivosmart 4 to participants who did not own compatible Garmin watches at the time of the data collection, while those who already owned one of the supported models used their own devices.

We shared the recruitment criteria and conditions on a webpage, and distributed our call for participants through press releases, social media, and mailing lists. We also directly contacted potential participants. A total of 301 applicants completed our sign-up questionnaire.

To ensure a comparable health profile across the three groups, additional selection criteria were applied:

1. No ongoing medical treatment or regular medication use.
2. Age between 25 and 65 years.
3. Smoking fewer than 20 cigarettes per day.
4. Consuming alcohol fewer than four times per week and less than 100 grams per drink.

This selection process resulted in 140 participants (meditation: 23, running: 57, control: 60) at the start of the study.

### Data Collection

We collected three types of data from three participant groups: physiological data from smartwatches, self-reported stress data through ESM, and questionnaires. We additionally obtained meditation start and end times for the meditation group.

We collected physiological and ESM data using our UTracker app, which leverages the Garmin Health SDK to capture beat-to-beat intervals and step counts from Garmin smartwatches, transmitting data to a server. Participants used this app to log data for any three weeks between October 12, 2022, and December 21, 2022. The app invoked ESM notifications randomly three times within their awake time, prompting participants to report their daily activity states (eating, work, housework, leisure, travel, rest, or other) and subjective stress on four levels (no, low, middle, or high stress). Responses submitted within 15 minutes were considered valid. Meditation group participants also recorded meditation session timings via an in-app feature.

After the three-week period, participants completed web-based questionnaires assessing physical activity, sleep quality, stress, mindfulness, and well-being. These included the International Physical Activity Questionnaire (IPAQ) [31], Pittsburgh Sleep Quality Index (PSQI) [32], Mindful Attention Awareness Scale (MAAS) [33], Perceived Stress Scale (PSS) [34], and WHO-5 Well-Being Index (WHO-5) [35], administered via LimeSurvey.

Ninety of our participants (meditation: 19, running: 32, control: 39) successfully reported at least two weeks of smartwatch and ESM data along with completed questionnaires. We therefore used the data from these 90 participants in the subsequent data analysis. The recruitment process, device usage, and demographic characteristics including group-wise distributions of sex, age, and BMI are summarized in the supplementary visualization (Multimedia Appendix 1).

### Calculation of HRV Indexes

We analyzed beat-to-beat intervals collected from smartwatches to calculate time-domain indices of heart rate variability (HRV) as objective markers of stress levels. These HRV indices were employed to compare HRV among the three groups and to evaluate the association of HRV with ESM-derived stress levels and meditation (hereafter, referred to as the comparative HRV analysis, the ESM-HRV analysis, and the meditation-HRV analysis, respectively).

Throughout all the three analyses, we employed the root mean square of successive differences (RMSSD), which is reported to strongly correlate with high-frequency HRV and be sensitive to parasympathetic activity changes [11]. To compute RMSSD, we divided the heart rate interval data into 5-minute non-overlapping windows, calculated the differences between successive intervals within each window, squared these differences, averaged the squared values, and then took the square root of the mean. In the comparative HRV analysis, 5-minute windows starting at 00:00 were employed. The ESM analysis focused on 5-minute windows centered around the reporting times, while the meditation analysis utilized 5-minute windows aligned with the reported start or end times.

For the comparative HRV analysis, we evaluated the distribution of mean RMSSD across the three groups. Mean RMSSD was calculated by averaging RMSSD values from all valid windows during the measurement period for each participant. To ensure the robustness of our findings, additional short-term HRV metrics, including SDNN, pNN50, and meanNN, were similarly assessed. Long-term HRV was also evaluated using the standard deviation of the normal-to-normal intervals (SDANN), calculated by averaging beat-to-beat intervals within 5-minute windows over a 24-hour period and determining the standard deviation of these means. SDANN provided a stable measure of long-term autonomic activity, complementing short-term metrics by mitigating the influence of transient fluctuations and artifacts [36].

To further investigate HRV differences while accounting for variations in physical activity levels across the groups, we stratified RMSSD by physical activity states. Using smartwatch-derived step counts and self-reported bedtime and wake-up time collected via the UTracker app, we categorized 5-minute windows into four states: run (≥400 steps), walk (50–399 steps), rest (10–49 steps or <10 steps outside sleep), and sleep (<10 steps during sleep). From windows assigned to each state, we calculated the mean RMSSD for each participant and compared the distributions among the three groups.

### Statistical Analysis

To compare questionnaire scores and HRV values across the three groups, we employed the Kruskal–Wallis test (K-W test). The pairwise comparisons were then performed using the Wilcoxon rank sum test (Wilcoxon test) combined with Shaffer’s multiple comparison procedure [37]. Statistical significance was defined as *P* < .05.

We tested associations of stress levels with daily activity states, using binomial tests on the EMS data. The four stress levels reported by the EMS responses were classified into two binary states: no/low and middle/high stress. Assuming the proportion of the EMS responses that reports the middle/high stress state in all the EMS reports as a binomial parameter, we tested the deviation of the middle/high stress proportion for each of the daily activity states. Similarly, we tested the deviation of the middle/high stress proportion for each of the meditation and running groups, assuming the middle/high stress proportion in the control group as a binomial parameter.

### Hierarchical Bayesian Analysis

In the ESM-HRV analysis, we examined differences in HRV across the two stress states, no/low and middle/high stress, by employing a hierarchical Bayesian model. The model was specifically designed to isolate HRV variations associated with stress states while accounting for inter-individual differences in baseline HRV (Multimedia Appendix 2). In the model, *m*, *i*, and *j* index stress states, participants, and observations, respectively. As the observed variable, the model has *mⱼ*, *iⱼ*, and *eⱼ*, which represent the stress state, participant, and RMSSD value in the *j*-th observation. The model assumes that *eⱼ* is sampled from a normal distribution with a mean of *βⱼ* defined as the sum of the mean RMSSD for the observed state (*αⱼ*) and the residual associated with the observed participant (*rⱼ*). By focusing on *αₘ* as a parameter corresponding to the mean RMSSD for state *m*, we estimated the posterior distributions of *αₘ*and the differences in *αₘ*by running Markov Chain Monte Carlo (MCMC).

Additionally, we extended the analysis to examine HRV differences between the two stress states within each of the three participant groups. For this purpose, the model was adapted such that m indexes six states, defined by the combination of the two stress levels and the three groups. In the meditation–HRV analysis, we examined HRV changes across the following meditation phases: the 10-minute before meditation phase, the during meditation phase, and the after meditation phases with 10-minute intervals up to 60 minutes post-meditation. Similarly, the model was adjusted to allow *m* to represent the respective meditation phases.

In the meditation–HRV analysis, we further investigated time-series HRV profiles across meditation practices of varying durations. For this purpose, we extended the model to assume temporal dependencies among parameters corresponding to mean RMSSD (Multimedia Appendix 3). In the model, *t*, *k*, *i*, and *j* index time points, meditation duration, participants, and observations, respectively. As the observed variable, the model has *tⱼ*, *kⱼ*, *iⱼ*, and *eⱼ*, which represent the time points, meditation duration, participant, and RMSSD value in the *j*-th observation. The parameter corresponding to the mean RMSSD for the time point *t* and the meditation length *k*, α*_t_^k^*, is sampled from a normal distribution with a mean of the parameter of the previous time point, α*_t-1_^k^*, though the parameter of the initial time point, α*_1_^k^*, depends on a hyper-parameter *α*. Taking the start or end of meditation as time 0, RMSSD was computed using start or end time-aligned windows spanning from –20 to +55 minutes. After grouping meditation practices by duration in 10-minute intervals, MCMC sampling was conducted to estimate posterior medians and 95% credible intervals of α*_t_^k^*, generating time-series HRV profiles for each duration group.

In all the analyses, MCMC sampling was performed on RStan version 2.32.6, with parameter settings including four chains, 1,000 burn-in iterations, and 2,000 total iterations. MCMC convergence was confirmed using the Gelman–Rubin diagnostic.

### Ethical Considerations

This study was approved by the Ethics Review Board of the Institute of Medical Science, The University of Tokyo (Approval Number: 2022-11-0629) and the Human Subject Research Ethics Review Committee of Tokyo Institute of Technology (Approval Number: 2022154). Participants provided informed consent by submitting their application after reading the study explanation on the recruitment website. Participation was voluntary, with withdrawal permitted at any time without justification.

As compensation, participants provided with Garmin smartwatches retained the devices after the data collection period. We offered the participants with compatible devices a Garmin electronic coupon.

## Results

### Questionnaire Data Analysis

We first compare subjective stress levels and related factors across the three groups, based on scores from the five questionnaires (Figure 2). The IPAQ results confirmed that the running group demonstrated significantly higher physical activity than the control group (medians: 2,625 and 990, respectively; K-W test: *P* < .001; Wilcoxon test with Shaffer correction: *P* < .001), with the meditation group scoring between the two (median: 1,548). In the MAAS questionnaire, the running group had significantly higher mindfulness scores than the control group (medians: 70.5 and 60, respectively; K-W: *P* = .006; Wilcoxon: *P* = .007). The meditation group also showed higher scores, although the difference did not reach the significance cutoff (median: 68; Wilcoxon: *P* = .099).

**Figure 2:**
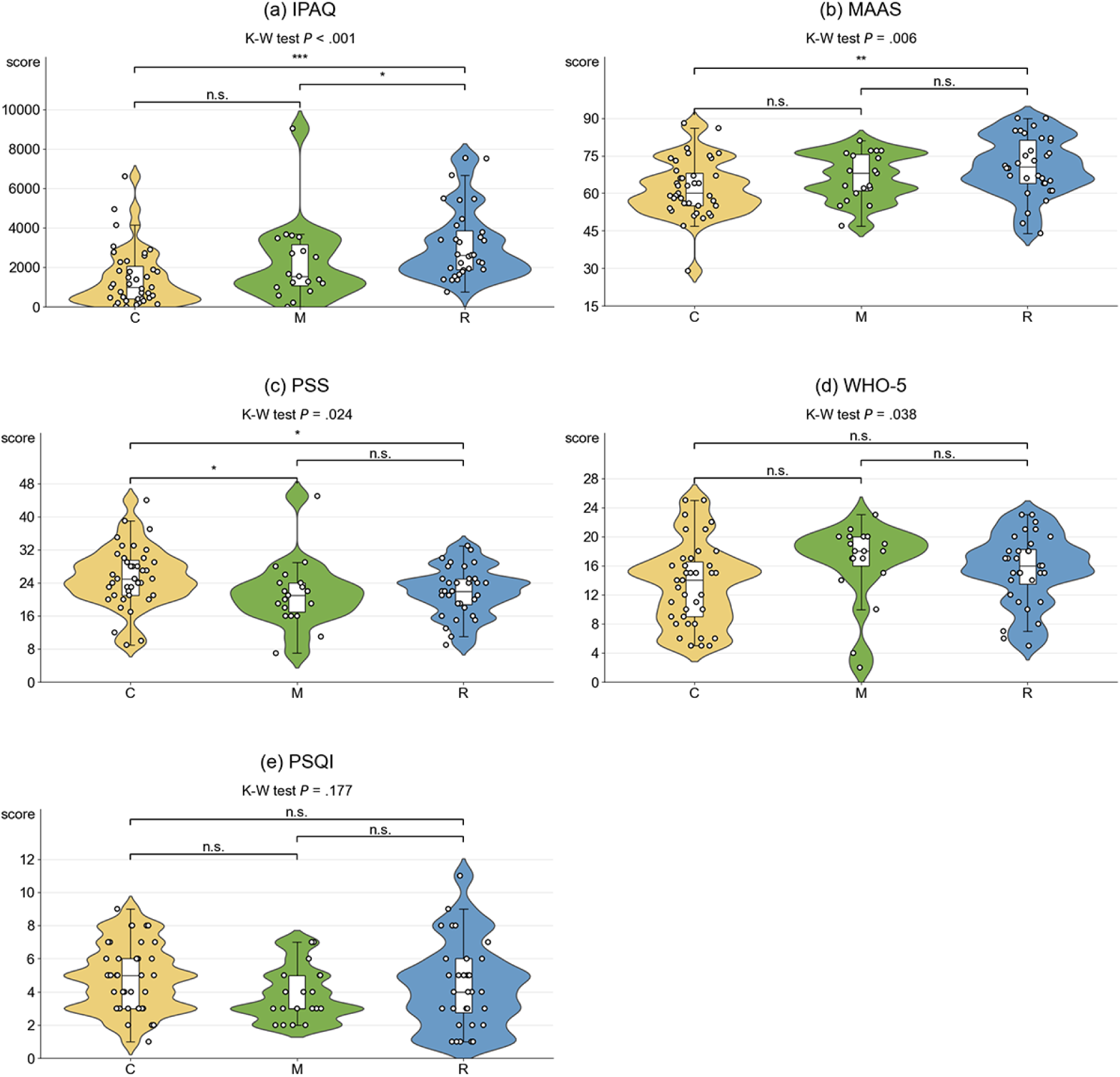
Comparative analysis of the five questionnaire scores. The horizontal line within each violin plot denotes the median, while the vertical line or shaded region represents the interquartile range, encompassing values from the 25th to 75th percentiles. Points represents scores from each participants in the control (*C*), meditation(*M*) and runner (*R*) groups. (a) high IPAQ scores indicate high physical activity levels, (b) high MAAS scores indicates high mindfulness levels, (c) low PSS scores low perceived stress levels, (d) high WHO-5 scores indicates high psychological well-being levels, and (e) low PSQI scores better sleep quality. Significant differences were assessed using K-W tests followed by Wilcoxon test. *n.s.* indicates not significant (*p >* 0.05), whereas the asterisks indicate significant difference (\*\*\**p <* 0.001, \*\**p <* 0.01, and \**p <* 0.05) in Wilcoxon test.

The PSS results indicate that both the meditation and running groups experienced significantly lower subjective stress compared to the control group (medians: 21[mediation], 22 [running] and 25[control]; K-W: *P* = .024; Wilcoxon: *P* = .048 and .040, respectively). In the WHO-5 questionnaire, a significant difference existed among the three groups (medians: 18[mediation], 16[running] and 14[control]; K-W: *P* = .038), and both the meditation and running groups showed higher well-being scores than the control group with marginal significance (Wilcoxon: *P* = .067 and .077, respectively). We observed no significant patterns in the PSQI questionnaire (medians: 3[mediation], 4[running] and 5[control]; K-W: *P* = .177).

These findings suggest that both meditation and physical exercise contributed to the reduction of stress together with enhanced mindfulness and well-being.

### HRV Data Analysis

We next compared smartwatch-derived HRV among the three groups (Figure 3a). Comparison of median RMSSD showed that the running group exhibited significantly higher RMSSD than the control group (medians: 47.016 and 42.017, respectively; K-W: *P* < .001; Wilcoxon: *P* < .001), In contrast, the meditation group exhibited RMSSD levels comparable to those of the control group (medians: 40.754 and 42.017, respectively). We also found a consistent pattern for other short-term HRV metrics (Multimedia Appendix 4), and SDANN, an HRV metric calculated over 24-hour periods (Figure 3b).

**Figure 3:**
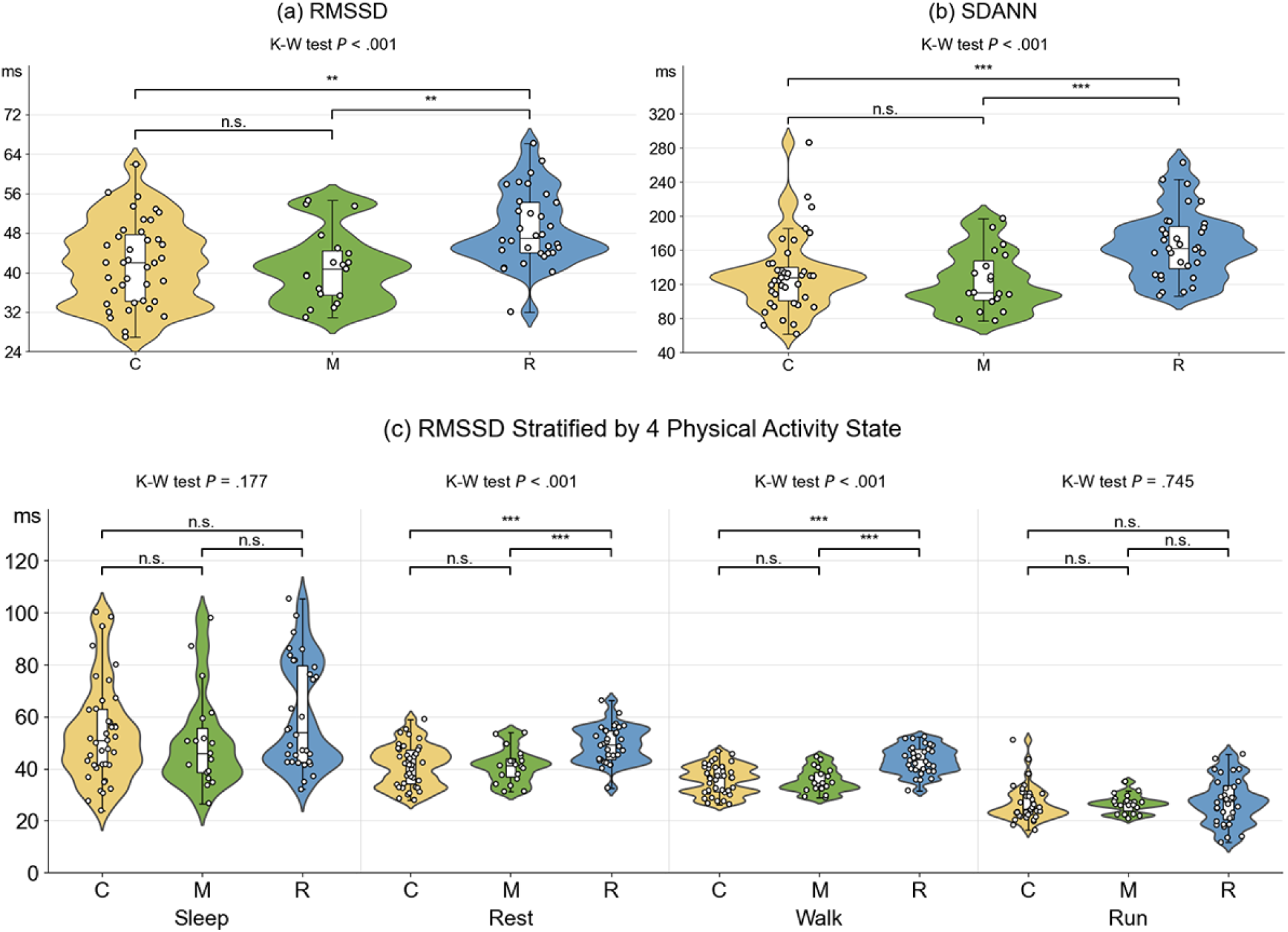
Comparative analysis of the HRV indexes. For each participant, the median values of various HRV indices were calculated from smartwatch-derived beat-to-beat intervals, and their distributions were compared across three groups: control (C), meditation (*M*), and runner (*R*). The indices include (a) 5-min RMSSD, (b) 24-hour SDANN, and (c) RMSSD stratified by four physical activity state (*sleep*, *rest*, *walk*, and *run*), categorized using smartwatch-derived step counts and self-reported sleep times. Figure presentation and statistical tests was conducted in the same manner as in Figure 2.

Since HRV is affected by the intensity of physical activity, it is possible that these results merely reflect the fact that the running group engaged in different physical activity intensities. To correct for such effects, we categorized 5-minute windows into four activity states—sleep, rest, walk, and run—using smartwatch step counts and self-reported sleep data. We calculated mean RMSSD for each activity state and compared group-level distributions (Figure 3c). The running group consistently displayed higher RMSSD during the rest state (medians: 49.150[running], 42.000[control], and 41.166[meditation]; K-W: *P* < .001; Wilcoxon: *P* < .001) and the walk state (medians: 43.451[running], 36.458[control], and 34.416[meditation]; K-W: *P* < .001; Wilcoxon: *P* < .001), compared to the other groups.

These results suggest that physical exercise is associated with higher HRV, while meditation practice has no apparent effects on overall HRV patterns in daily life.

### ESM Data Analysis

In this study, we also collected daily activity states and stress levels by ESM to analyze their associations (Figure 4a). ESM samplings were randomly administered three times per day, generating 4,557 responses (51.78 per participant on average; Multimedia Appendix 5). Among all the responses, 10.3% reported middle/high stress. Using this proportion as a baseline, stress levels were higher during housework (14.0%; *P* = .024, based on a binomial test) and work (20.7%; *P* < .001), and lower during leisure (2.3%; *P* < .001), rest (2.9%; *P* < .001), and eating (3.4%; *P* < .001). This pattern was consistent across groups, although the proportion of middle/high stress responses was lower in the running group (7.2%; *P* < .001) and meditation group (8.6%; *P* < .001) compared to the control group (13.7%), in line with the questionnaire findings. We also examined the relationship between self-reported stress levels and RMSSD recorded at reporting times (Figure 4b). To account for inter-participant variations in baseline HRV, we employed a hierarchical Bayesian approach to estimate posterior distributions of the average RMSSD for the two different stress states: no/low stress and middle/high stress (Multimedia Appendix 2).

**Figure 4:**
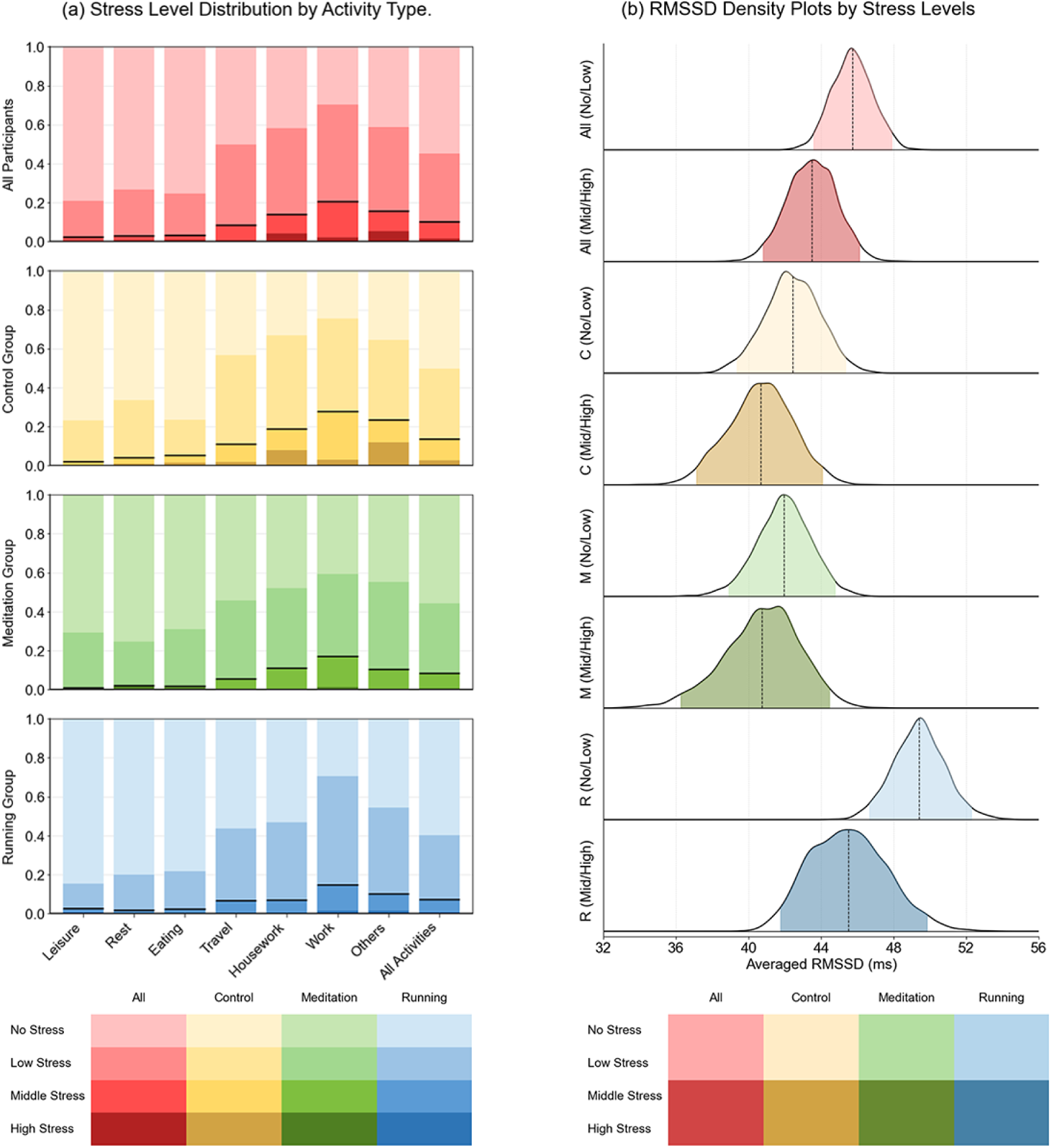
Associations with Stress Levels with Daily Activities and HRV. (a) The proportion of self-reported stress levels across seven daily activity states, derived from 4557 ESM responses. The vertical axis denotes the proportions of four-staged stress levels, while the horizontal axis represents activity types. Horizontal black lines across the bars indicate the proportion of *no/low stress* conditions. (b) Posterior density plots of averaged RMSSD under *no/low stress* and *middle/high stress* conditions. RMSSD was calculated using 5-minute windows centered on reporting times. A hierarchical Bayesian model (Figure 8) was employed to account for variability in response frequency and baseline HRV across participants. Posterior densities of *α_m_* were estimated using MCMC, with shaded regions denoting 95% credible intervals and vertical dotted lines indicating posterior medians.

The analysis revealed that RMSSD was lower during middle/high stress states compared to no/low stress states, confirming the ecological validity of HRV as a stress indicator. We additionally sampled the posterior distribution of the difference between the two parameters, whose posterior median estimated the RMSSD difference as −2.242 (95% credible interval: −3.967 to −0.260; Multimedia Appendix 6). This trend was observed across all groups, with the running group showing a more pronounced decrease of −3.939 ms (95% credible interval: −7.041 to −0.737), likely due to higher baseline RMSSD values.

Collectively, our ESM analysis demonstrated that subjective stress levels are associated with both daily activity states and HRV.

### Analysis of HRV and Meditation

From the meditation group, we collected information about start and end times of 632 daily meditation practices (33 per participant on average) using the smartphone app. We investigated HRV changes during meditation by integrating the meditation timing information with the heartbeat data obtained from smartwatches (Multimedia Appendix 7).

From each participant in the meditation group, RMSSD was computed for the 10-minute period before meditation, the period throughout meditation, and the periods in 10-minute intervals up to 60 minutes after meditation. Similarly to the ESM data analysis, we employed a hierarchical Bayesian model to account for inter-participant variations in baseline HRV and calculate posterior distributions of parameters corresponding to averaged RMSSD values for each period (Figure 5a). As a result, RMSSD showed an increase during meditation, with the posterior median estimated at 4.675 (95% CI: 2.962–6.380; Multimedia Appendix 8). Remarkably, RMSSD required more than 30 minutes to return to baseline levels after meditation, suggesting a prolonged residual effect.

**Figure 5:**
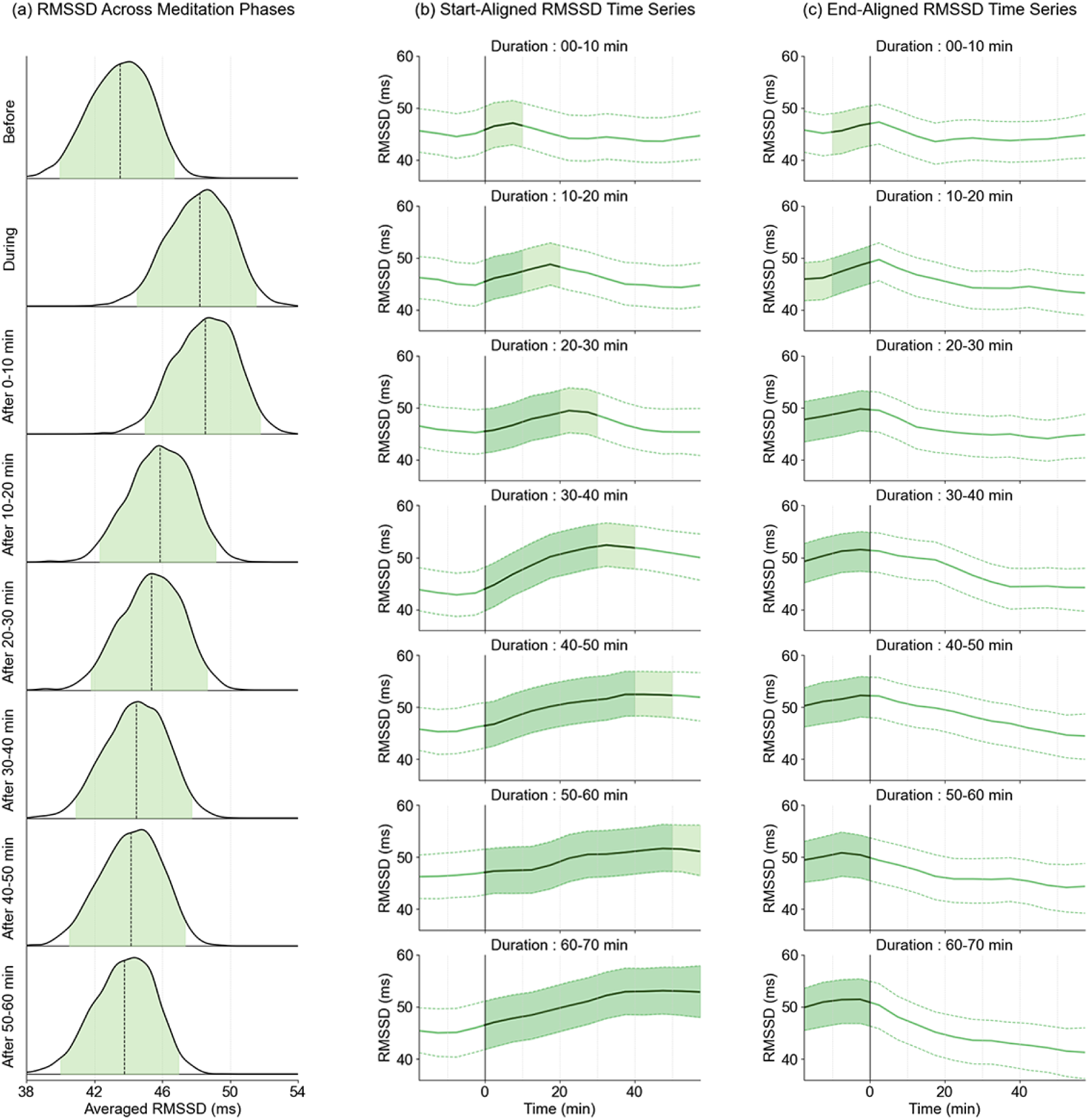
Variation of HRV levels during daily mediation practices. (a) Posterior density plots illustrating the averaged RMSSD across different phases of meditation practice. RMSSD data were categorized as follows: the 10-minute *before* meditation phase, the *during* meditation phase, and the *after* meditation phases with 10-minute intervals up to 60 minutes post-meditation. RMSSD was computed using start time-aligned 5-minute windows for the *before* and *during* phases, while end time-aligned windows were employed for the *after* phases. The data were analyzed and visualized as in Figure 4 b. (b) Time-series RMSSD profiles across meditation practices of varying durations. Assuming the start of meditation as time 0, RMSSD was calculated using start time-aligned windows spanning from *−*20 to +55 minutes. Profiles were grouped by meditation duration in 10-minute intervals, and MCMC-based hierarchical Bayesian analysis (in Figure 8) was used to estimate posterior medians and 95 % credible intervals of *α^k^*, represented by solid and dotted lines, respectively. Horizontal time scales were aligned with the centers of the RMSSD windows. The shaded regions between the dotted lines denote the *during* meditation phase, while the lightly shaded areas indicate that the results are based on data partially originating from outside the *during* meditation phase. (c) Time-series RMSSD profiles assuming the end of meditation as time 0 and basing on end time-aligned windows, visualized in the same manner as b.

We also examined RMSSD profiles across meditation practices of varying durations. Meditation practices were stratified into 10-minute intervals based on their duration, and time-series RMSSD trajectories were computed using a hierarchical Bayesian approach (Figure 5b). The analysis revealed a progressive increase in RMSSD that persisted throughout the meditation period. Upon stopping meditation, RMSSD gradually decreased toward baseline levels, underscoring the persistence of residual effects.

In summary, our analysis revealed that during daily meditation practice, practitioners experience an increase in HRV, which may persist for a few tens of minutes after meditation.

## Discussion

In this study, we used mHealth technology to examine the relationship between stress and HRV in meditation practitioners, recreational runners, and a control group without regular mental or physical exercise routines. Questionnaires revealed significantly lower stress levels in both meditation practitioners and recreational runners, supporting established evidence of stress reduction through meditation and exercise. However, smartwatch-derived HRV data showed elevated HRV in recreational runners while meditation practitioners exhibited HRV levels comparable to the control group. Although multiple intervention studies have assessed the impact of meditation practice on HRV, their findings have been mixed; some report increases in daily-life HRV among meditation groups relative to controls, while others observe no significant differences [21–25]. In our observational study involving experienced meditation practitioners, we found no evidence of a general increase in HRV throughout daily life.

Using a smartphone-based experience sampling method (ESM) integrated with smartwatch-derived heart rate monitoring, we investigated the association between stress and HRV across three groups. ESM also captured daily activities, allowing us to examine the links between stress and activity states. Across all groups, elevated stress levels were consistently associated with reduced real-time HRV, as well as with specific stress-related activities such as work. Consistent with the questionnaires, ESM confirmed that both meditation and running groups experienced lower stress levels in daily life compared to controls. While recreational runners showed more pronounced stress-related reductions in HRV, likely due to elevated baseline levels, meditation practitioners and controls exhibited comparatively smaller shifts. Meditation training has been shown to attenuate HRV reductions during standardized cognitive stress tasks [38]. Taken together with this, our findings suggest that meditation practitioners may perceive identical stress-inducing conditions as less stressful than non-practitioners, resulting in smaller reductions in HRV. Although the absence of overall group differences in daily-life HRV may appear inconsistent with this interpretation, a plausible explanation is that real-world stressors intense enough to elicit marked HRV changes arise only infrequently in naturalistic settings.

We also examined how HRV changes during daily meditation practice by integrating HRV data with meditation timing information collected via a smartphone app. Our analysis demonstrated a significant HRV increase during meditation, which persisted for a few tens of minutes afterward, suggesting a substantial residual effect. To our knowledge, such a residual HRV elevation after meditation has not been previously documented. By capturing meditation practices in a naturalistic, daily-life context, this study provides unique insights into the physiological dynamics of meditation. Meditation practitioners’ ability to increase HRV at an arbitrary timing, along with the prolonged residual effect, may correlate with stress reduction; however, this hypothesis warrants further investigation with adequate controls.

While most HRV studies have relied on ECG-based devices, we utilized a Garmin smartwatch with PPG sensors, enabling the collection of three weeks of continuous data. Although the reliability of PPG-derived HRV metrics has been debated due to susceptibility to noise [13–14], the combination of extensive data collection and robust statistical approaches allowed us to extract meaningful physiological insights. Especially, the hierarchical Bayesian approach used in our ESM–HRV and meditation–HRV analyses proved effective in isolating signals associated with covariates of interest while accounting for inter-participant variability in baseline HRV. Aligned with a recently published work [39], our achievement highlights the potential of Bayesian approaches to enhance the utility of wearable PPG-based devices in HRV research.

Despite these promising results, our study remains preliminary, with several limitations requiring attention. For instance, participant group composition introduced potential biases. A substantial proportion of the running group used their own Garmin smartwatches for data collection, while the control and meditation groups primarily used study-provided devices (Multimedia Appendix 1). This discrepancy arose from recruiting runners via Garmin users’ mailing list. This could influence the results due to differences in device models and the users’ familiarity with the devices, although we confirmed that recreational runners wearing different device models showed no significant difference in HRV levels (data not shown).

The meditation group also harbored potential recruitment biases. Participants in the meditation group were recruited through calls on social media and direct outreach to potential candidates. In the recruitment process, we noticed that more experienced meditation practitioners were often reluctant to wear a smartwatch on a daily basis, which may have biased our sample toward individuals with less meditation experience. Additionally, the meditation group included participants practicing different meditation styles, which may have introduced heterogeneity. In the questionnaire analysis, the mindfulness scores showed no significant difference, which may reflect not only the small sample size but also these concerns related to participant recruitment.

It is also important to consider the possibility that the three groups had covariates that could influence our analyses (Multimedia Appendix 1). For example, while HRV is known to vary with age, the age distributions differed across the groups.

Additionally, we observed variations in BMI distributions and sex proportions among the groups. Future studies should recruit a sufficient number of participants to account for these covariates through covariance analysis.

Despite these limitations, our findings underscore the potential of mHealth technology to transform meditation research by enabling real-time assessment of physiological changes, such as HRV, in naturalistic daily-life settings. The methodology developed in this study provides a foundation for both large-scale observational studies and intervention studies to directly measure the effects of meditation. Furthermore, our approach, which captures the residual effects of meditation, would offer a promising tool to evaluate the proficiency and impact of meditation, paving the way for personalized and effective wellness strategies.

## Acknowledgements

We thank Yuki Ooishi, NTT Communication Science Laboratories, for insightful discussion about HRV analysis.

## Data Availability

The datasets generated and analyzed during this study are available from the corresponding author on reasonable request.

## Authors’ Contributions

J.T.: data processing and analysis, and manuscript drafting. S.G.: app development, and data collection. M.F.: study design, participant recruitment, and data analysis supervision. M.M.: participant recruitment. K.S.: data analysis supervision. K.Y.: app development supervision. A.N.: project management, study design, participant recruitment, data collection, data analysis, and manuscript refinement.

## Conflicts of Interest

None declared.

## Abbreviations

mHealth: Mobile Health
HRV: Heart Rate Variability
ESM: Experience Sampling Method
MBSR: Mindfulness-Based Stress Reduction
MBCT: Mindfulness-Based Cognitive Therapy
IPAQ: International Physical Activity Questionnaire
PSQI: Pittsburgh Sleep Quality Index
MAAS: Mindful Attention Awareness Scale
PSS: Perceived Stress Scale
WHO-5: WHO-5 Well-being Index
RMSSD: Root Mean Square of Successive Differences
SDNN: Standard Deviation of NN Intervals
pNN50: Percentage of Adjacent NN Intervals Greater Than 50 ms
meanNN: Mean of NN Intervals
SDANN: Standard Deviation of All 5-Minute Mean NN Intervals
MCMC: Markov Chain Monte Carlo

---

**Multimedia Appendix 1:**
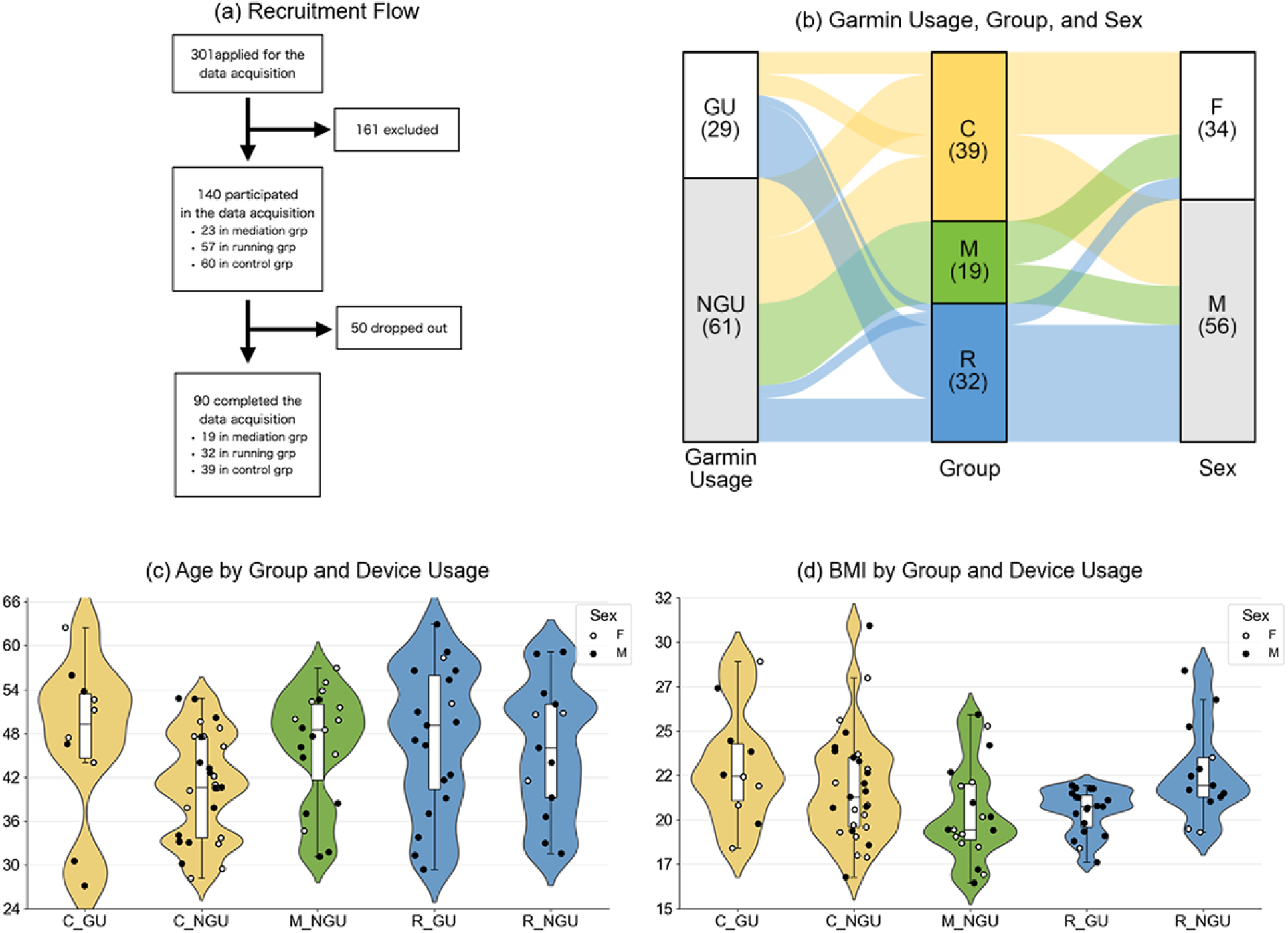
Characteristics of the recruited participants. (a) Recruitment flow chart of participants assigned to three groups: meditation (*M*), running (*R*), and control (*C*). (b) Sankey diagram illustrating the relationships between Garmin usage, group classification, and sex distribution. GU represents Garmin users, who participated in the study with their own smartwatches, while NGU represents non-Garmin users, who participated using provided smartwatches. The width of the flows corresponds to the number of participants transitioning between categories, with numbers in parentheses indicating the total participants in each category. (c) Age and (d) Body Mass Index (BMI) distribution across the three groups and differences in Garmin usage. In the violin plots (as presented in Figure 2), white dots represent female participants, while black dots represent male participants. BMI was calculated as weight (kg) divided by the square of height (m²).

**Multimedia Appendix 2:**
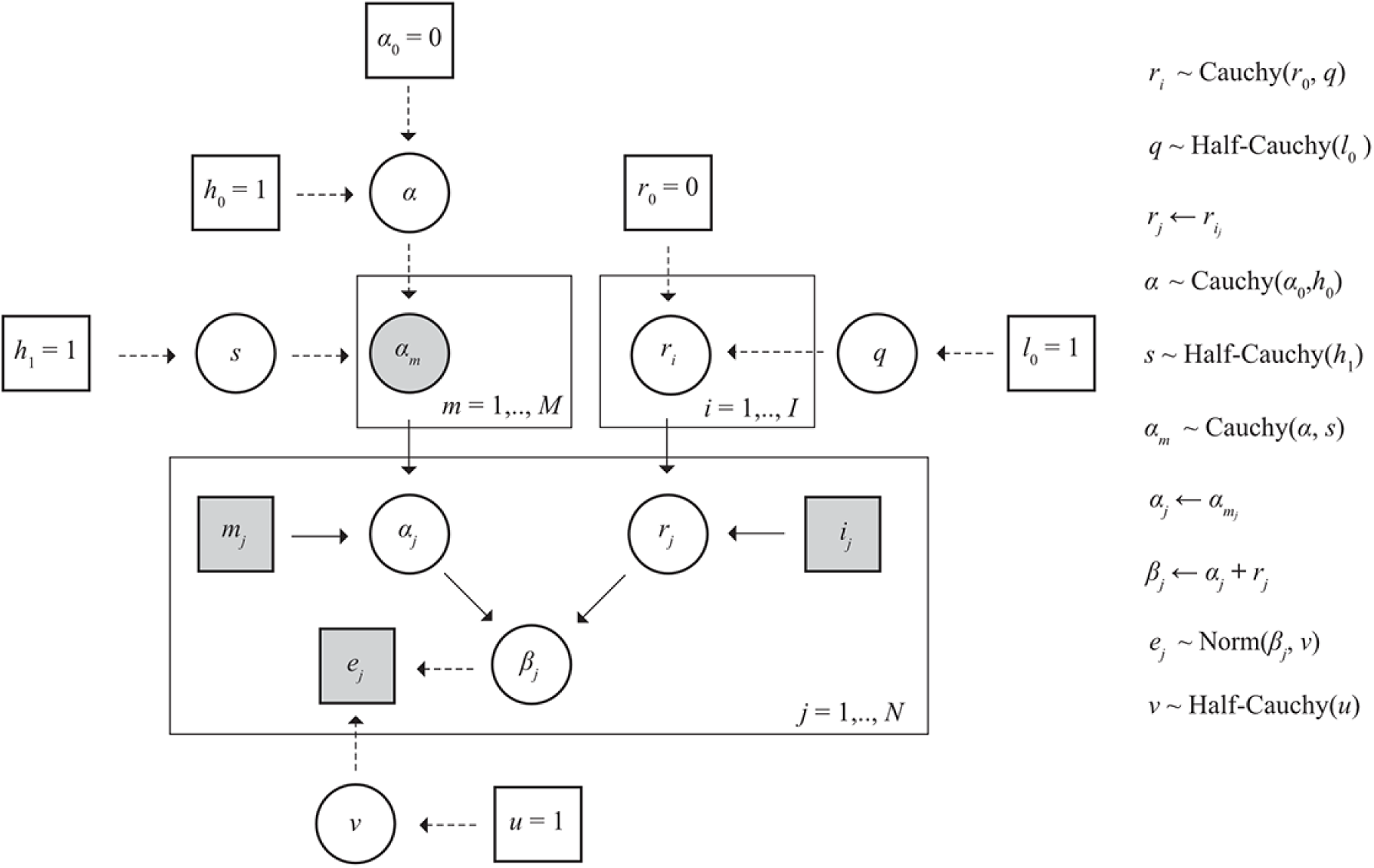
Comparative analysis of short-term HRV indexes. Violin plots illustrating comparisons of three metrics across groups. (a) SDNN denotes the standard deviation of beat-to-beat intervals, (b) pNN50 represents the percentage of successive beat-to-beat intervals differing by more than 50 ms, and (c) meanNN indicates the average of beat-to-bea intervals over 5-minute intervals. Figure presentation and statistical tests was conducted in the same manner as in Figure 2.

**Multimedia Appendix 3:**
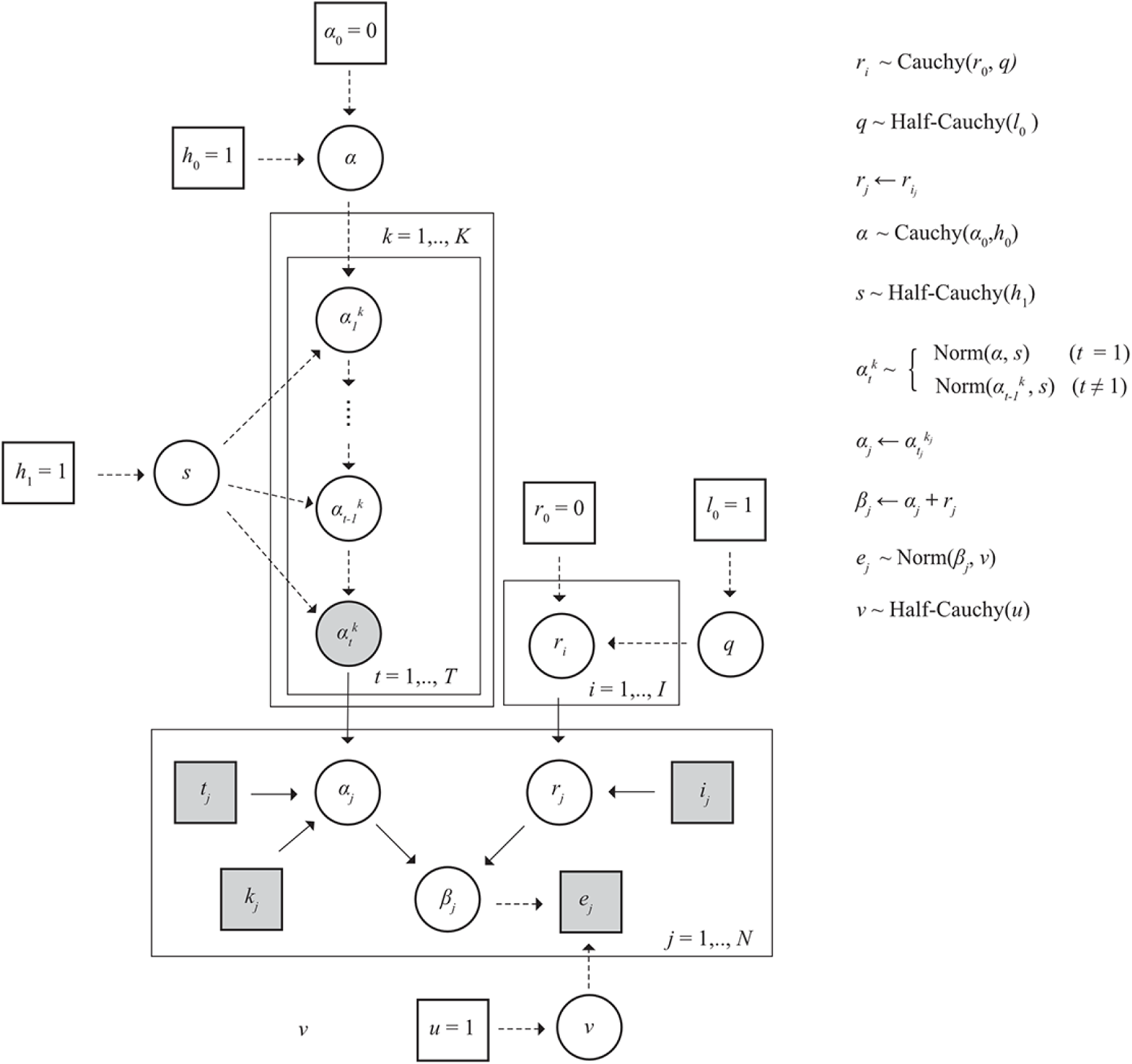
Probabilistic graphical representation of the model used for the comparison of RMSSD between stress levels or meditation phases. Circles and rectangles represent random variables and fixed constants, respectively. Shaded squares indicate observed data while a shaded circle indicates a variable of interest. Solid and dashed arrows represent probabilistic and deterministic dependencies, respectively. For definition of variables, see Methods.

**Multimedia Appendix 4:**
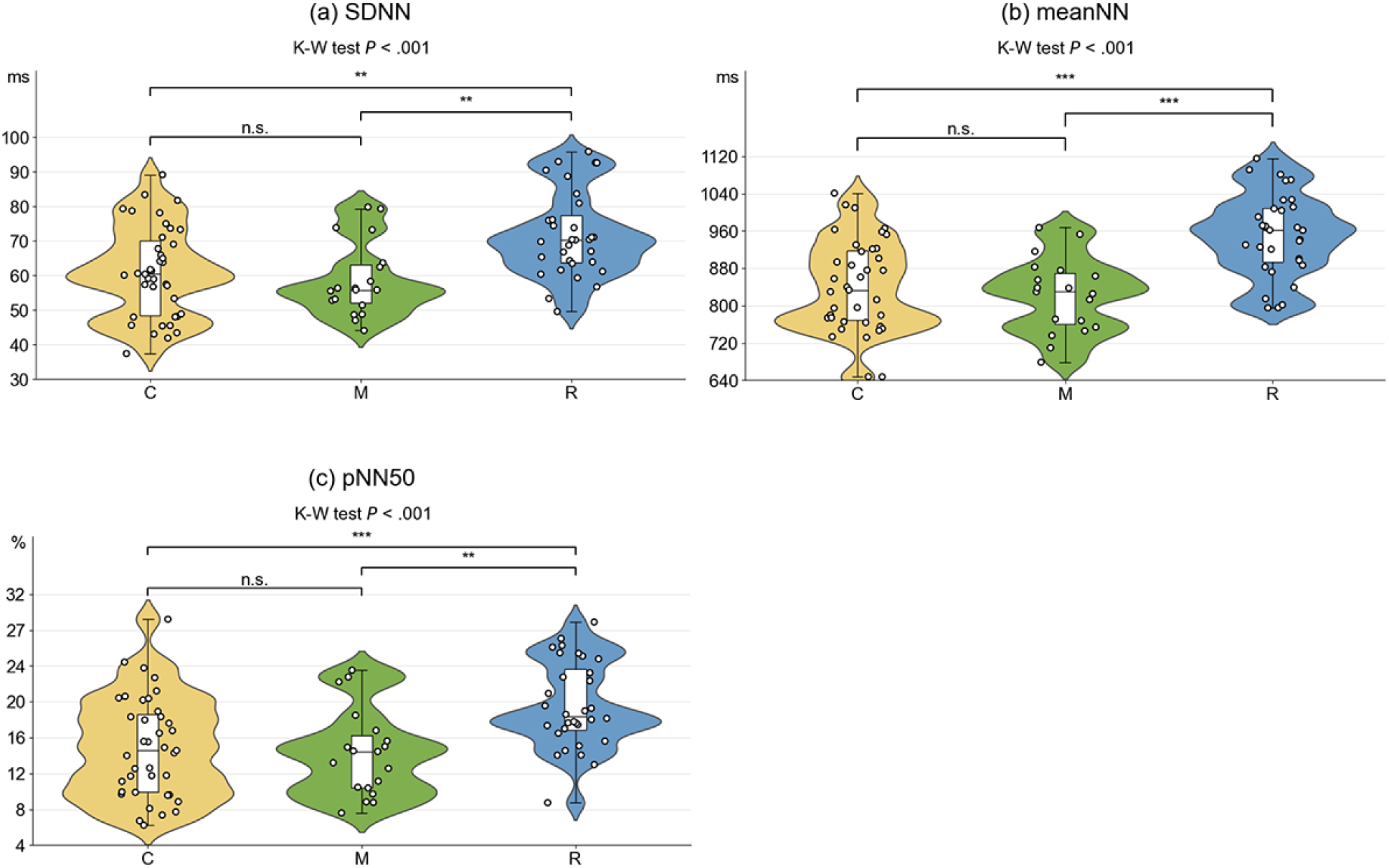
Probabilistic graphical representation of the model used for the time-series analysis of RMSSD along meditation practices. Presented as in Supplementary Figure 8.

**Multimedia Appendix 5:**
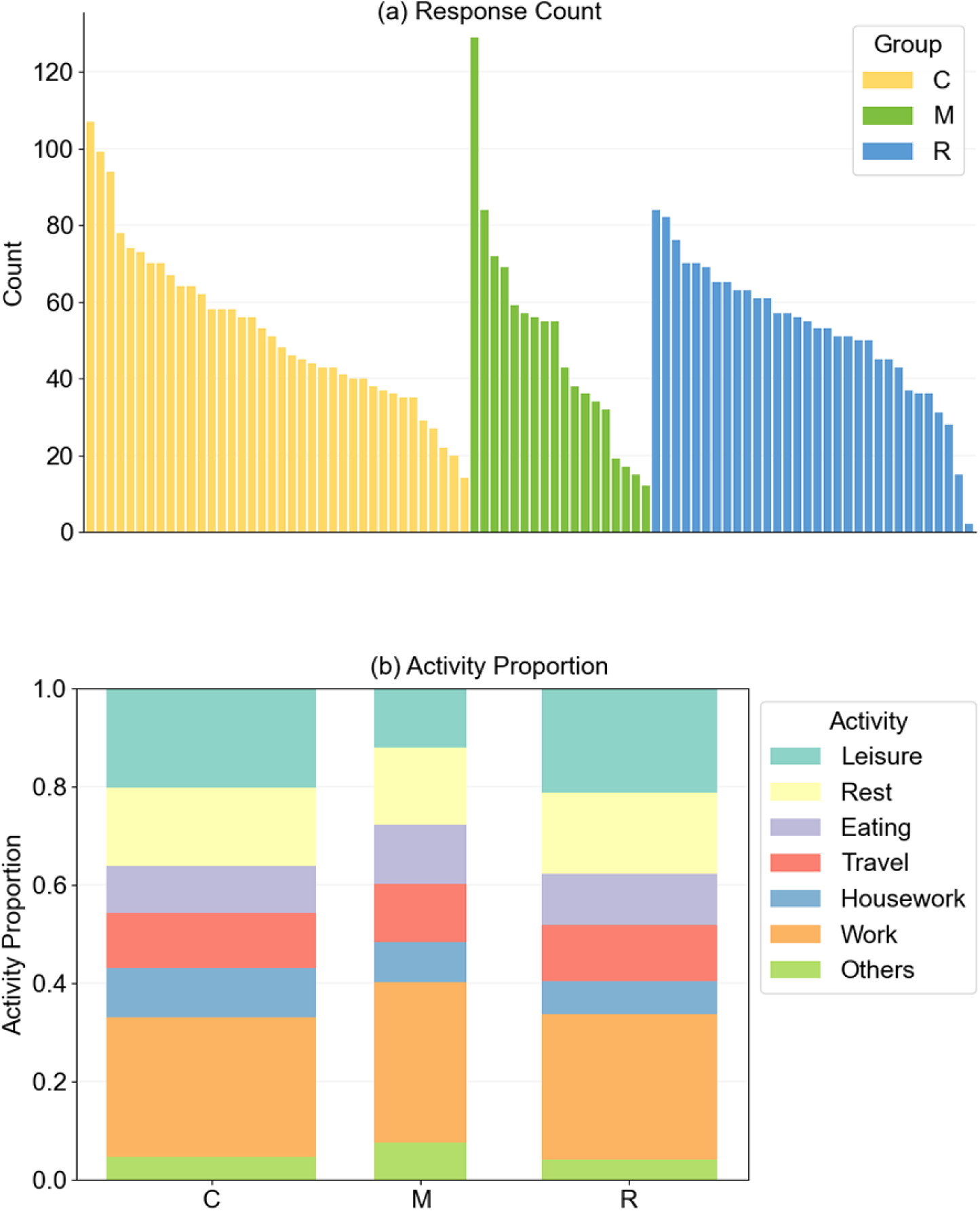
Overviews of the ESM data. (a) A bar charts showing the counts of ESM responses across participants. Bar height reflects the total number of responses for each participant. Groups are presented from left to right as control (*C*), meditation (*M*), and running (*R*), with participants within each group ordered by the number of responses in descending order. (b) A stacked bar chart showing the proportions of seven dairy activity states for each of the three groups. Bar widths are scaled according to the total number of ESM responses from participants in each group.

**Multimedia Appendix 6:**
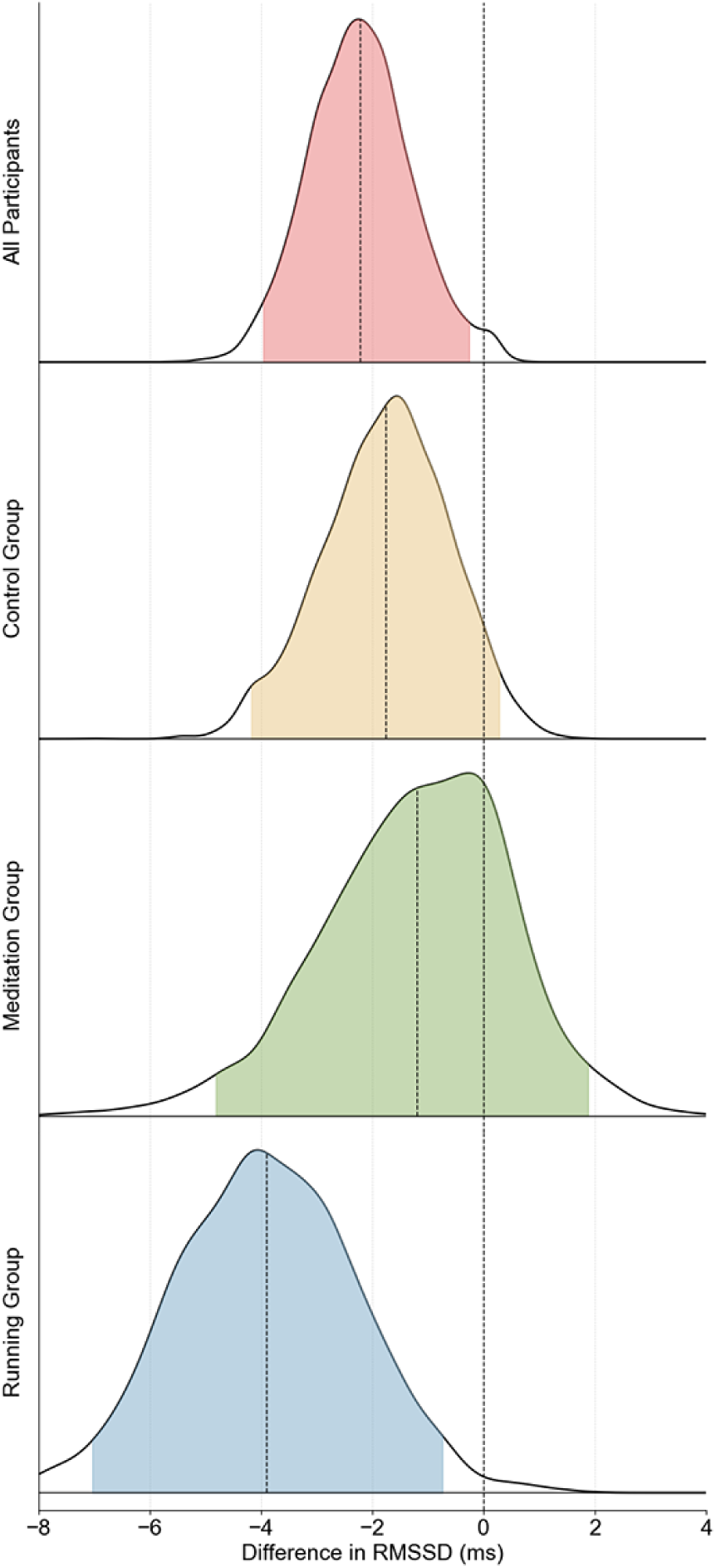
Posterior density plots of differences in averaged RMSSD at high stress conditions with reference to low stress conditions. In the MCMC sampling process used to generate Figure 4 (b), posterior distributions were simultaneously estimated for differences in parameters corresponding to averaged RMSSD (*α_m_*) between the different stress levels. The posterior densities are presented as in Figure 4 (b). The vertical highlighted dotted line indicates no difference, and negative value on the horizontal axis indicates a reduction in RMSSD under *middle/high stress* compared to *no/low stress*.

**Multimedia Appendix 7:**
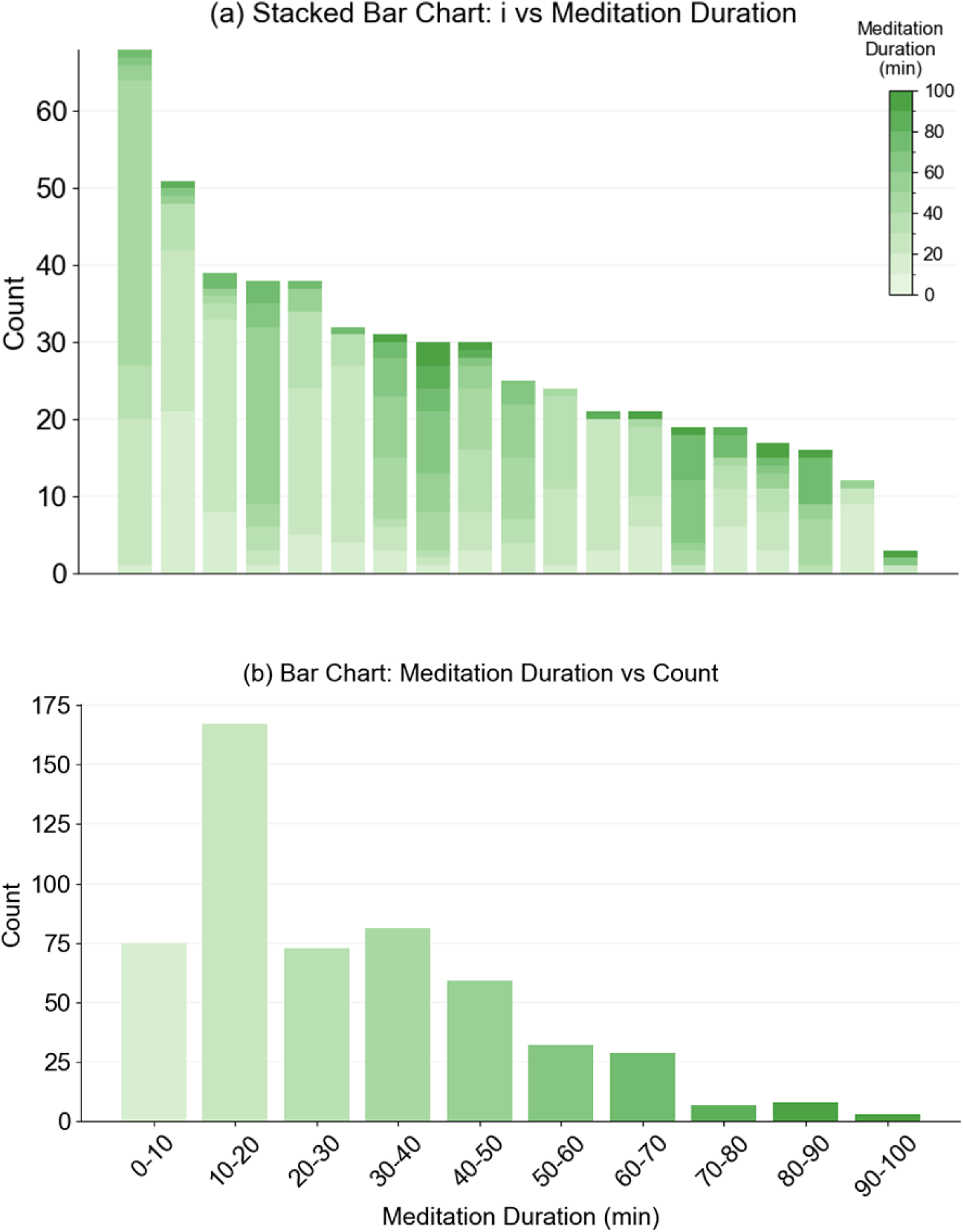
Overviews of the meditation records. (a) A stacked bar chart showing the number of meditation practices recorded by participants in the meditation groups. Each bar is composed of stacked segments, with each segment representing a single meditation practice from an individual participant. The height of each bar reflects the total count of meditation practices, with participants arranged in descending order along the horizontal axis based on their total count. Within each bar, stacked segments are also ordered in descending duration, with darker shades representing longer meditation practices. (b) A bar chart showing the distribution of meditation practice durations. Practices recorded by all the meditation group members are categorized into 10-minute intervals based on their duration, with the height of each bar representing the total number of practices whose durations fall within each interval. Bar colors correspond to the coding scheme shown in (a).

**Multimedia Appendix 8:**
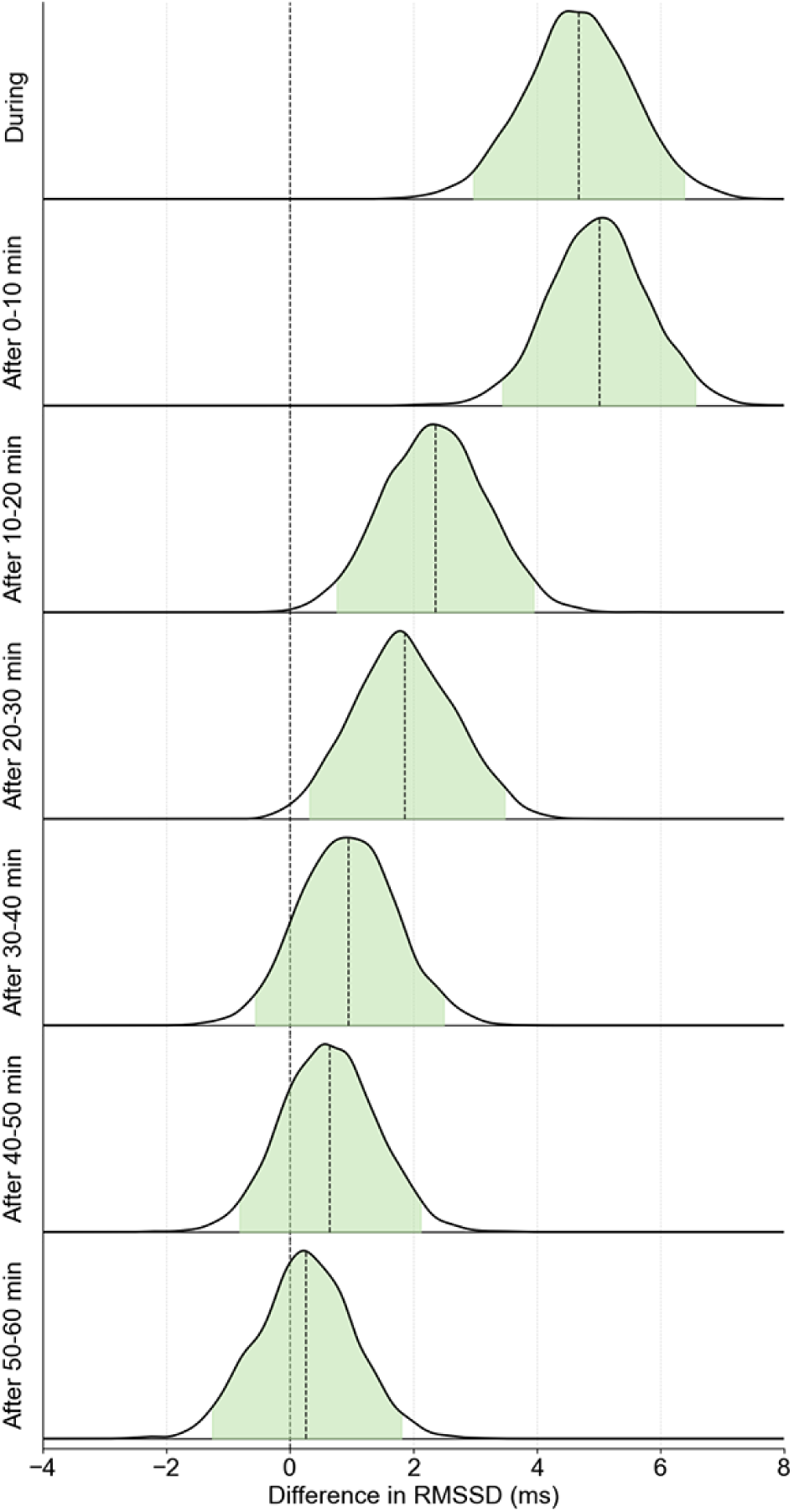
Posterior density plots of differences in averaged RMSSD at each meditation phase with reference to the “Before” meditation phase. In the same manner as in Figure 11, posterior density plots of parameters *α_m_*were generated from the MCMC sampling process used to generate Figure 5 (a). A positive value on the horizontal axis indicates an increase in RMSSD under each meditation phase compared to the *before* meditation phase.

## Notes

### Competing Interest Statement

The authors have declared no competing interest.

### Funding Statement

This study did not receive any specific grant from funding agencies in the public, commercial, or not-for-profit sectors. Garmin Japan Ltd. provided electronic coupons to some participants as part of their compensation for participation.

### Author Declarations

Ethics Review Board of the Institute of Medical Science, The University of Tokyo gave ethical approval for this work (Approval Number: 2022-11-0629). Human Subject Research Ethics Review Committee of Tokyo Institute of Technology gave ethical approval for this work (Approval Number: 2022154).

